# The epidemiological investigation of vitamin D deficiency in northern Henan province

**DOI:** 10.1101/2023.09.11.23295345

**Authors:** Huiling Deng, Ziyang Lin, Junzheng Yang

**Affiliations:** The Third Affiliated Hospital of Xinxiang Medical University, Department of clinical laboratory, Xinxiang 453000, China; Guangdong nephrotic drug Engineering Technology Research Center, Institute of Consun Co. for Chinese Medicine in Kidney Diseases, Guangdong Consun Pharmaceutical Group, Guangzhou 510530, China

**Keywords:** vitamin D, investigation, epidemiology, prevention, northern Henan province

## Abstract

**Aim/purpose:** To analyze the epidemiology of vitamin D deficiency in northern Henan province, to lay the foundation for prevention and treatment of vitamin D deficiency and the vitamin D related diseases.

**Methods:** We collected the basic information including gender, age, and vitamin D concentration of 22184 people who detected the 25 hydroxyvitamin D concentration in the Third Affiliated Hospital of Xinxiang Medical University from June 2020 to May 2023, analyzed the vitamin D deficiency distribution in male children population, female children population, male adult population and female adult population; and the differences of 25 hydroxyvitamin D concentration of diabetes mellitus patients in the total investigation population in northern Henan province were also analyzed and compared according to gender, age and different seasons.

**Results:** A total of 22184 data were collected from June 2020 to May 2023 in the Third Affiliated Hospital of Xinxiang Medical University; the age range of investigation objects was from 1 month to 93 years old; there were 8176 male people and 14008 female people, accounting for 36.86% and 63.14% in the total investigation population, respectively; there were 1318 diabetes mellitus patients, accounting for 5.94% in the total investigation population; there were 796 males and 522 females, accounting for 60.39% and 39.61% in the total 1318 diabetes mellitus patients, respectively; the investigation results demonstrated that the number of people with vitamin deficiency was 278, the number of people with vitamin insufficiency was 1418, and the number of people with normal vitamin concentration was 20488, accounting for 1.25%, 6.39% and 92.35% in the total investigation population, respectively; in the adult female population, the number of people with vitamin deficiency and with vitamin insufficiency were highest compared the male children population, female population and male adult population; the statistical results demonstrated that there were the significantly statistical differences among vitamin D concentration<25nmol/mL population, vitamin D concentration at 25-49 nmol/mL population, and vitamin D concentration at 50∼80 nmol/mL in female adult population at the different age (P=0.0039); and there were no statistical differences in male children population, male adult population and female children population at different ages and at different vitamin D concentrations (P>0.05); the results demonstrated that there were the significantly differences among vitamin D concentration<25nmol/mL population, vitamin D concentration at 25-49 nmol/mL population, and vitamin D concentration at 50∼80 nmol/mL in adult diabetes mellitus population at gender, age and different seasons.

92.35% people in northern Henan province had the normal vitamin concentration, there were 7.65% people with vitamin insufficiency or vitamin deficiency in northern Henan province; the number of people with vitamin deficiency and with vitamin insufficiency were highest in the adult female population, and there were the significantly statistical differences in female adult population at the different age at different vitamin concentration population and in diabetes mellitus adult population at gender, age and different seasons.

**Conclusion:** The incidence rate of vitamin insufficiency or vitamin deficiency in northern Henan province was 7.65%, and the epidemiology of vitamin D deficiency in northern Henan province had the specific characteristics. Those evidences may provide useful information for prevention and treatment of vitamin D deficiency and vitamin D related diseases.

## Introduction

Vitamin D is a lipid soluble cyclopentane polyhydrophenanthrene compound as a kind of special organic active regulators which is indispensable to the human body by regulating calcium and phosphorus metabolism [1,2], promoting bone growth [3], regulating cell growth and differentiation [4,5], and regulating immune response [6,7]. Many evidences demonstrated that vitamin D deficiency could result in many kinds of diseases including schizophrenia, cardiovascular diseases, metabolic diseases, and immune diseases, and it is reported that about 7% of people suffered from severe vitamin D deficiency, especially in some countries in the Middle East, North Africa, and Asian countries in the world, there are about one-third of people have serum 25 hydroxyvitamin D concentration below normal standards [8,9], which may cause the huge economic burden no matter for the country but also for individuals. Therefore, conducting the nutritional status evaluation to understand the local vitamin D deficiency situation and investigate the epidemiology of vitamin D is very necessary to understand vitamin D deficiency in different population and at different age for prevention and treatment of vitamin D deficiency. For this purpose, we collected the basic information of 22184 people who detected the 25 hydroxyvitamin D concentrations in the Third Affiliated Hospital of Xinxiang Medical University from June 2020 to May 2023 to investigate the epidemiology of vitamin D deficiency in northern Henan province, hope to provide some useful information for local medical staff.

## Materials and Methods

A total of 22184 data including gender, age and vitamin D concentration who detected the 25 hydroxyvitamin D concentration were collected in the Third Affiliated Hospital of Xinxiang Medical University from June 2020 to May 2023; vitamin D concentration was measured by chemiluminescence immunoassay. Those data were classified and analyzed according to gender, age and vitamin D concentration; the differences of 25 hydroxyvitamin D concentrations in diabetes mellitus patients in northern Henan province were also analyzed and compared according to gender, age and seasons. Evaluation standard of vitamin D nutritional status was as following: vitamin D sufficiency: vitamin D concentration between 50-80 nmol/mL; vitamin D insufficiency: vitamin D concentration between 25-49 nmol/mL; vitamin D deficiency: vitamin D concentration <25 nmol/mL.

## Results

In the total of 22184 data, there were 8176 male people and 14008 female people, accounting for 36.86% and 63.14% in the total investigation population, respectively; there were 5418 people in the male children population, 3717 people in the female children population, 2758 people in the male adult population, 10291 people in the female adult population; accounting for 24.42%, 16.76%, 12.43% and 46.39% in the total investigation population, respectively; the investigation results demonstrated that the number of people with vitamin deficiency was 278, the number of people with vitamin insufficiency was 1418, and the number of people with normal vitamin concentration was 20488, accounting for 1.25%, 6.39% and 92.35% in the total investigation population, respectively; in the adult female population, the number of people with vitamin insufficiency and with vitamin insufficiency were highest compared the male children population, female population and male adult population; the statistical results demonstrated that there were the significantly statistical differences among vitamin D concentration<25nmol/mL population, vitamin D concentration at 25-49 nmol/mL population, and vitamin D concentration between 50∼80 nmol/mL in female adult population at the different age (P=0.0039); and there were no statistical differences in male children population, male adult population and female children population at different ages and at different vitamin D concentrations (P>0.05) (Table 1).

**Table 1.**
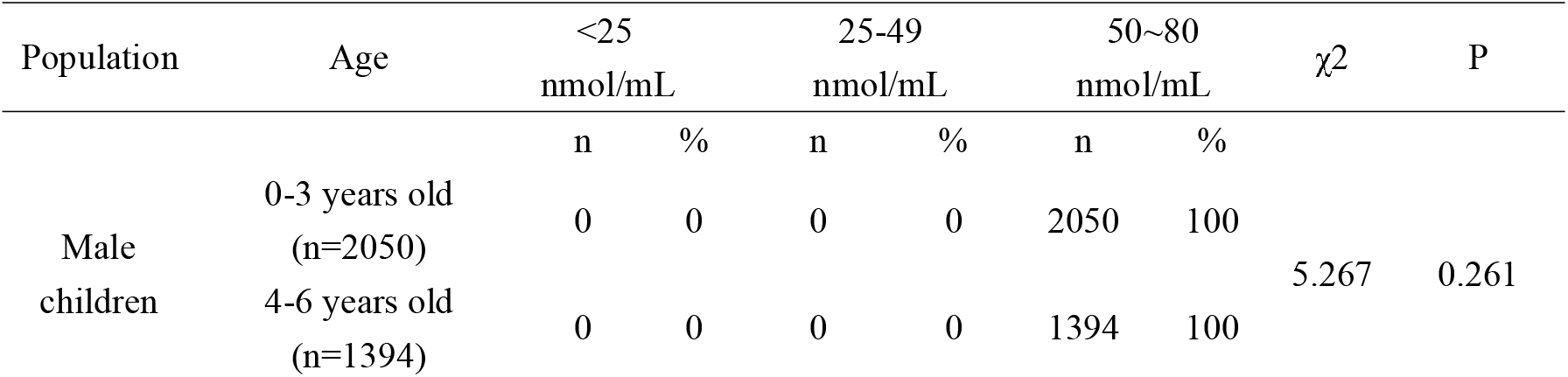
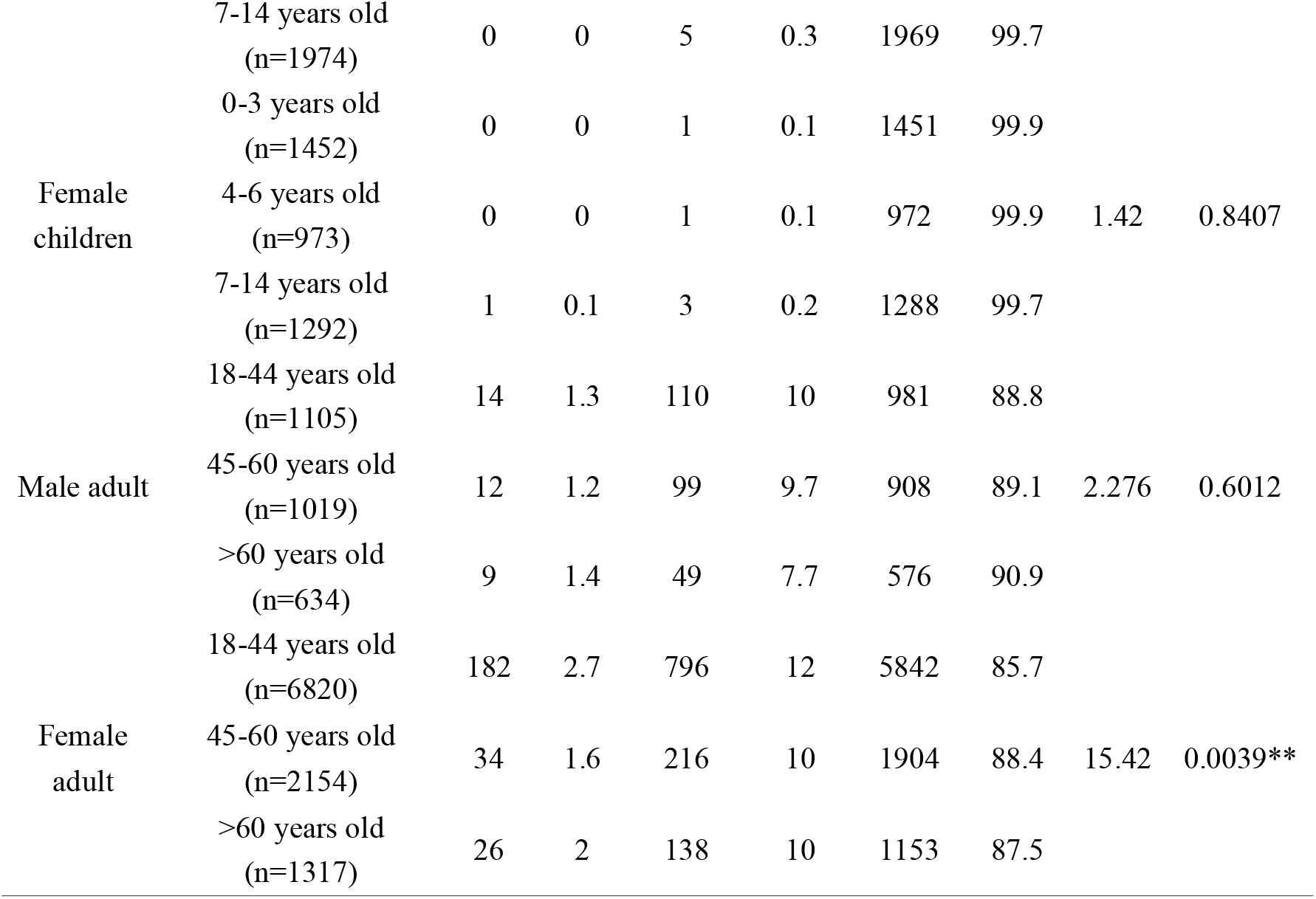
The distribution of vitamin D concentration in different population in northern Henan province.

**Table 2.**
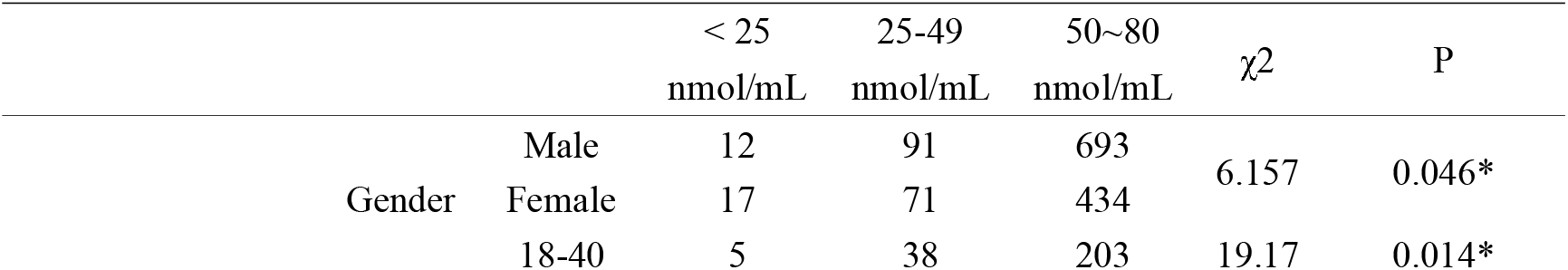
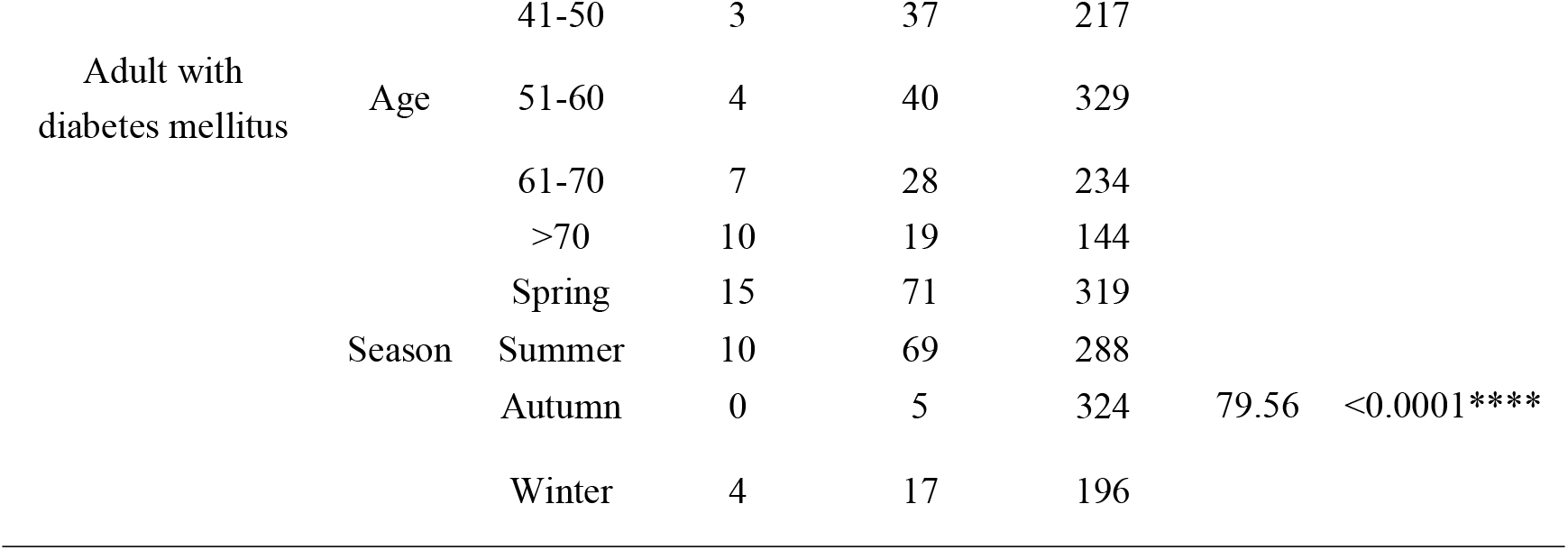
The epidemiology of vitamin D deficiency in adult population with diabetes mellitus in northern Henan province.

| | | < 25<br>nmol/mL | 25-49<br>nmol/mL | 50~80<br>nmol/mL | $\chi^2$ | P |
| --- | --- | --- | --- | --- | --- | --- |
| Gender | Male | 12 | 91 | 693 | 6.157 | 0.046* |
|  | Female | 17 | 71 | 434 |  |  |
|  | 18-40 | 5 | 38 | 203 | 19.17 | 0.014* |

|  |  |  |  |  |  |  |  |
| --- | --- | --- | --- | --- | --- | --- | --- |
| Adult with<br>diabetes mellitus | Age | 41-50 | 3 | 37 | 217 |  |  |
|  |  | 51-60 | 4 | 40 | 329 |  |  |
|  |  | 61-70 | 7 | 28 | 234 |  |  |
|  |  | >70 | 10 | 19 | 144 |  |  |
|  | Season | Spring | 15 | 71 | 319 |  |  |
|  |  | Summer | 10 | 69 | 288 |  |  |
|  |  | Autumn | 0 | 5 | 324 | 79.56 | <0.0001**** |
|  |  | Winter | 4 | 17 | 196 |  |  |

And then, we analyzed the distribution of vitamin D concentration in adult diabetes mellitus population in northern Henan province. The results demonstrated that a total of 1318 adult with diabetes mellitus were collected, there were 796 males and 522 females, accounting for 60.39% and 39.61% in adult diabetes mellitus population, respectively; the age range was from 18-93 years old; the results demonstrated that there were the significantly differences among vitamin D concentration<25nmol/mL population, vitamin D concentration at 25-49 nmol/mL population, and vitamin D concentration at 50∼80 nmol/mL population in adult diabetes mellitus population at gender (P=0.046), age (P=0.014) and seasons (P<0.0001).

## Discussion

Vitamin D plays the important role in regulating body metabolism and maintaining normal physiological functions in the human body, vitamin D deficiency or vitamin D insufficiency could cause a series of diseases including osteomalacia [10], rheumatoid arthritis [11], diabetes mellitus [12], Crohn’s disease [13] and immune diseases [14]; seasonal emotional disorders are also associated with vitamin D deficiency [15], different age stages including pregnancy and early childhood growth also require vitamin D supplementation [16,17]. The evidences demonstrated that people with limited outdoor activities, vegetarians, patients with celiac disease, people taking some kinds of special medications, and other risk factors including high latitude, lack of sunlight, age, obesity also could result in the vitamin D deficiency [18]. Therefore, study on the epidemiological vitamin D and understanding the distribution characteristics of vitamin D deficiency among different populations is of great significance for prevention and reduction of vitamin D deficiency and vitamin D related diseases.

In our article, 22184 vitamin D data were collected from June 2020 to May 2023 in the Third Affiliated Hospital of Xinxiang Medical University, found that there were 7.65% people with vitamin D insufficiency or vitamin D deficiency in northern Henan province, the proportion of vitamin D insufficiency or vitamin D deficiency in adult female population was 82.08%, the data demonstrated that the incidence rate of vitamin D insufficiency or vitamin D deficiency in Henan province was relatively low; vitamin D insufficiency or vitamin D deficiency mainly occurred in adult female population, especially in adult female population aged at 18-44 years old (n=6820), therefore, This type of population requires special attention, and the necessary measures to take to prevent or treat the proportion in adult female population.

Diabetes mellitus is a kind of chronic diseases which occurs when the pancreas does not produce enough insulin or the body cannot effectively utilize the insulin produced, the growing evidences demonstrated that the vitamin D deficiency was associated with the progression of diabetes mellitus [19, 20, 21]. Especially, Wan Z, et al. demonstrated that the higher concentration of serum 25 (OH) D in vivo was significantly associated with the lower risk of cardiovascular complications in type 2 diabetes patients, and the risk tends to be stable when 25 (OH) D reaches 50 nmol/L [22]. In our data, we also found that the relationship between the vitamin D deficiency and diabetes mellitus in the north Henan province; and a total of 1318 adult with diabetes mellitus were collected in our investigation, the statistical results demonstrated that there were the significantly differences among vitamin D concentration<25nmol/mL population, vitamin D concentration at 25-49 nmol/mL population, and vitamin D concentration at 50∼80 nmol/mL population in diabetes mellitus adult population at gender (P=0.046), age (P=0.014) and seasons (P<0.0001). those results demonstrated that the vitamin D deficiency may affect the situation of the adult diabetes mellitus patients, and gender, age, and seasons could be the main risk factors.

## Conclusion

The incidence rate of vitamin D deficiency in north Henan Province was relatively low, and the epidemiology of vitamin D deficiency in north Henan Province had the specificity. Those results may provide some useful information for the prevention and treatment for vitamin deficiency and vitamin related diseases.

## Human and animal rights

Not applicable.

## Data Availability

All data produced in the present work are contained in the manuscript

## Acknowledgements

None.

